# Progression / remission of Coronavirus disease 2019: Data driven recommendations for repeating SARS-CoV-2 nucleic acid amplification tests

**DOI:** 10.1101/2020.06.16.20132001

**Authors:** Kaitlyn Williams, Reed Idriss, Jessica Dodge, Samuel Barasch

## Abstract

**Aims:** This short study was performed to better understand the time frame associated with changes in SARS-CoV-2 nucleic acid testing and provide recommendations for repeat testing. Recommendations were useful as little guidance is available for repeat testing in patients being followed expectantly for changes in disease.

**Methods:** A review of laboratory data of tests for SARS-CoV-2 nucleic acid was performed selecting patients who had changing results. Time between changes in test results was determined to provide guidance for repeat testing.

**Results:** The interquartile range of data for patients who had a negative to positive change in lab testing (“progression”) was 6-16 days (Median 9). The interquartile range of data for patients who had a positive to negative change in test results (“remission”) was 9-21 days (Median 14).

**Conclusion:** Because sampling of the nares or nasopharynx can be variable, repeat testing should be performed swiftly when symptomatic patients are negative. The data in this short study varies widely, so authors recommend repeat testing during a period of time associated with the interquartile range or median (see results above).

## INTRODUCTION

Laboratory testing for Coronavirus disease 2019 (COVID-19) has taken a prominent position in the SARS-CoV-2 pandemic, and nucleic acid testing has been negative in some symptomatic patients^1^ and positive in some patients who are asymptomatic.^1-3^ Results which are discordant from the clinical picture are likely due to a combination of several factors including the following: viral prodromal/incubation period^1,4,5^, sampling discordance from different swabbing techniques and clinical abilities^6^, analytical differences between nucleic acid detection / instruments/methods or other technical analytical complications^6,7^, and changes in patient disease state^1,2^. Given that analytical sensitivity is high with nucleic acid amplification (testing methodology available early in the pandemic), clinical and biological factors such as appropriate specimen collection technique, progression of disease, and regression of disease can contribute to changes in values for the diagnostic nucleic amplification for SARS-CoV-2. This study was undertaken to assess prevalence of laboratory data changes in SARS-CoV-2 nucleic acid amplification tests, as well as time frames associated with these changes. Findings could guide testing for progression or remission of disease and may contribute to biological knowledge of the disease.

## METHODS

This study was reviewed by our institutional review board and received exempt determination. The laboratory information system was searched for SARS-CoV-2 nucleic acid amplification tests in inpatients and outpatients associated with Western Connecticut Health Network (WCHN) hospitals (Danbury, CT; Norwalk, CT; New Milford, CT) through testing performed at WCHN laboratories, Connecticut Department of Public Health Laboratory, and Reference Laboratories between 3/1/2020 and 4/19/2020. All tests were based upon presence of nucleic acid for SARS-CoV-2 in the patient nasopharynx or nasal cavity. Testing laboratory, test manufacturer, and viral gene targets include the following: WCHN, Abbott ID NOW™, RdRP gene; Connecticut Department of Public Health Laboratory, CDC primers, N1/N2/N3 genes; Sunrise Medical Laboratories, Roche Cobas 6800/8800, ORF1 and E genes.^8-10^ For testing performed on site at hospitals, testing was performed according to manufacturer specifications. Prior to 4/6/2020, all samples were collected via nasopharyngeal swabs or nasal swabs and stored/transported to testing facilities in Universal Transport Media (UTM). Beginning on 4/6/2020, nasal swabs without UTM were tested on Abbott ID NOW™ analyzers. Each test was performed on a separate sample. Patients with duplicate testing were identified. From the group of patients with duplicate testing, a group of patients with discordant results was identified. Result verified date was used to assign chronology. Number of days between discordant results was determined. Patients changing from a negative to positive result were classified as “progression” while patients changing from positive to negative were classified as “remission.” “Progression” and “Remission” in this study refer to newly appearing positive results or newly appearing negative results. In this context, the terms “progression” and “remission” imply a change of laboratory testing from negative to positive or positive to negative. The number of days between the first positive or negative result and the first discordant result was recorded as the number of days to progression or remission. Patients with same day results which were discordant were recorded as 0 days between progression or remission. Figure 1 histogram was created with Microsoft Excel. R 4.0.0 was used to generate scatterplots and boxplots and statistical analysis.

**Figure 1:**
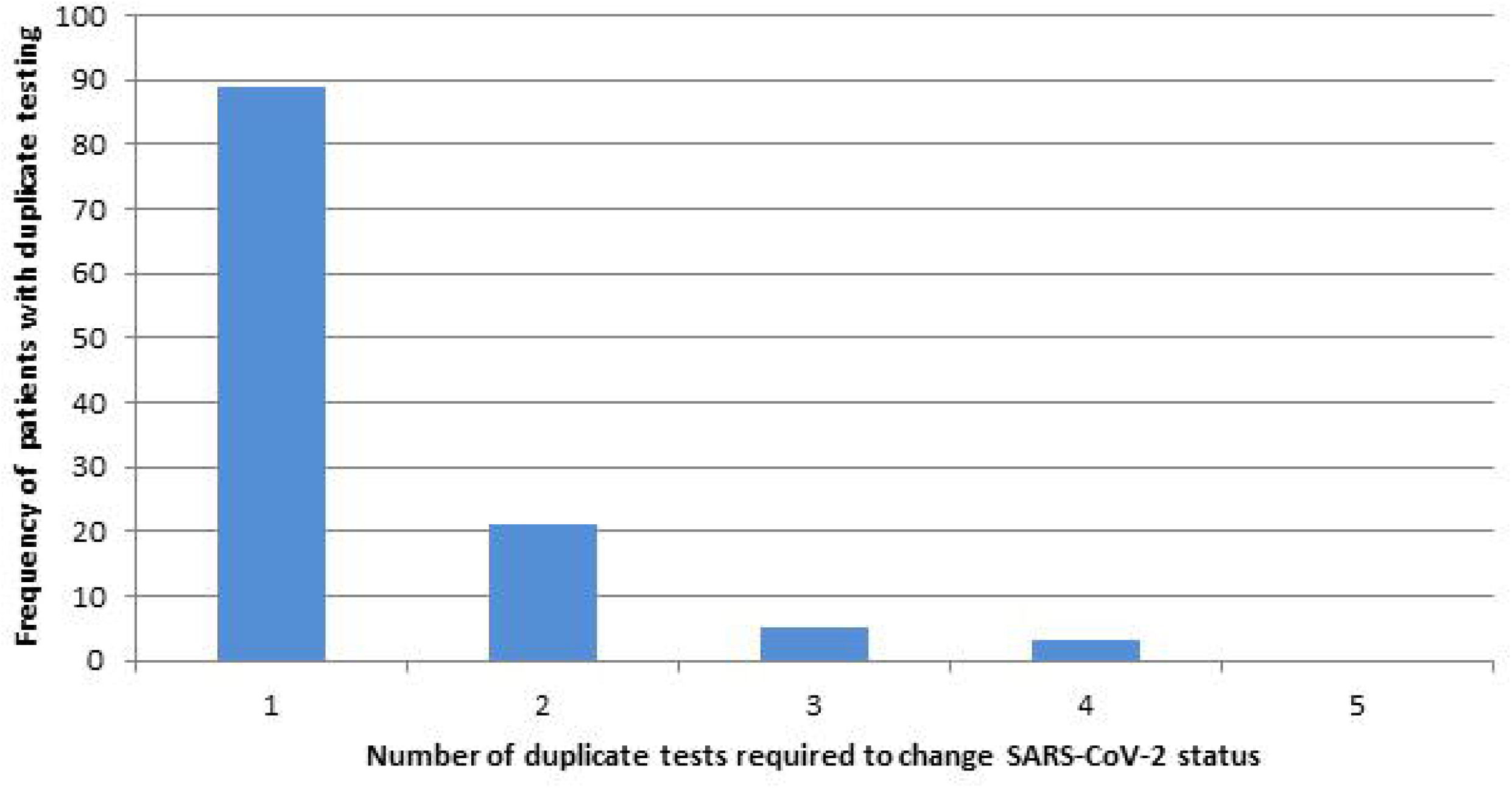
Repeated tests necessary for change in SARS-CoV-2 detected status.

## RESULTS

**Table 1:** Summary of results

|  |  |
| --- | --- |
| Number of nucleic acid amplification tests for SARS-CoV-2 in study time period | 13,206 |
| Number of patients tested | 12,360 |
| Number of patients with more than one test | 709 |
| Number of patients with more than one test and no changes in test results | 591 |
| Number of patients with a change in test result | 118 |
| Number (percentage) of patients with a change in test result on 1 or 2 or tests | 89 (75%) – patients with change after 1 test.<br>110 (93%) – patients with change after 2 tests. |
| Number of patients with Progression (change from negative to positive) | 65 |
| Number of patients with Regression (change from positive to negative) | 53 |
| Interquartile range (Median) Progression | 6-16 days (9 days) |
| Interquartile range (Median) Regression | 9-21 days (14 days) |

13,206 nucleic acid amplification tests were performed and resulted in the study time period. Most laboratory result dates were between 0 and 2 days following the order date with order date being shortly before date and time of specimen collection. The total number of tests represented 12,360 patients tested. 591 patients had 688 duplicate tests which were concordant. 118 patients had 158 discordant duplicate SARS-CoV-2 tests. 65 patients with discordant tests were classified as “progression” while 53 patients with discordant tests were classified as “regression”. 3 patients had 0 days to progression and 1 patient had 0 days to remission. Most patients with change in laboratory values had a change after 1 test (Figure1). For patients with “progression” the mean number of days was 10.4 (10.9 excluding days to progression of 0) and the median was 9 (Figure 2). For patients with “remission” the mean was 13.5 days (13.7 excluding days to remission of 0) and median was 14 days (Figure 3).

**Figure 2:**
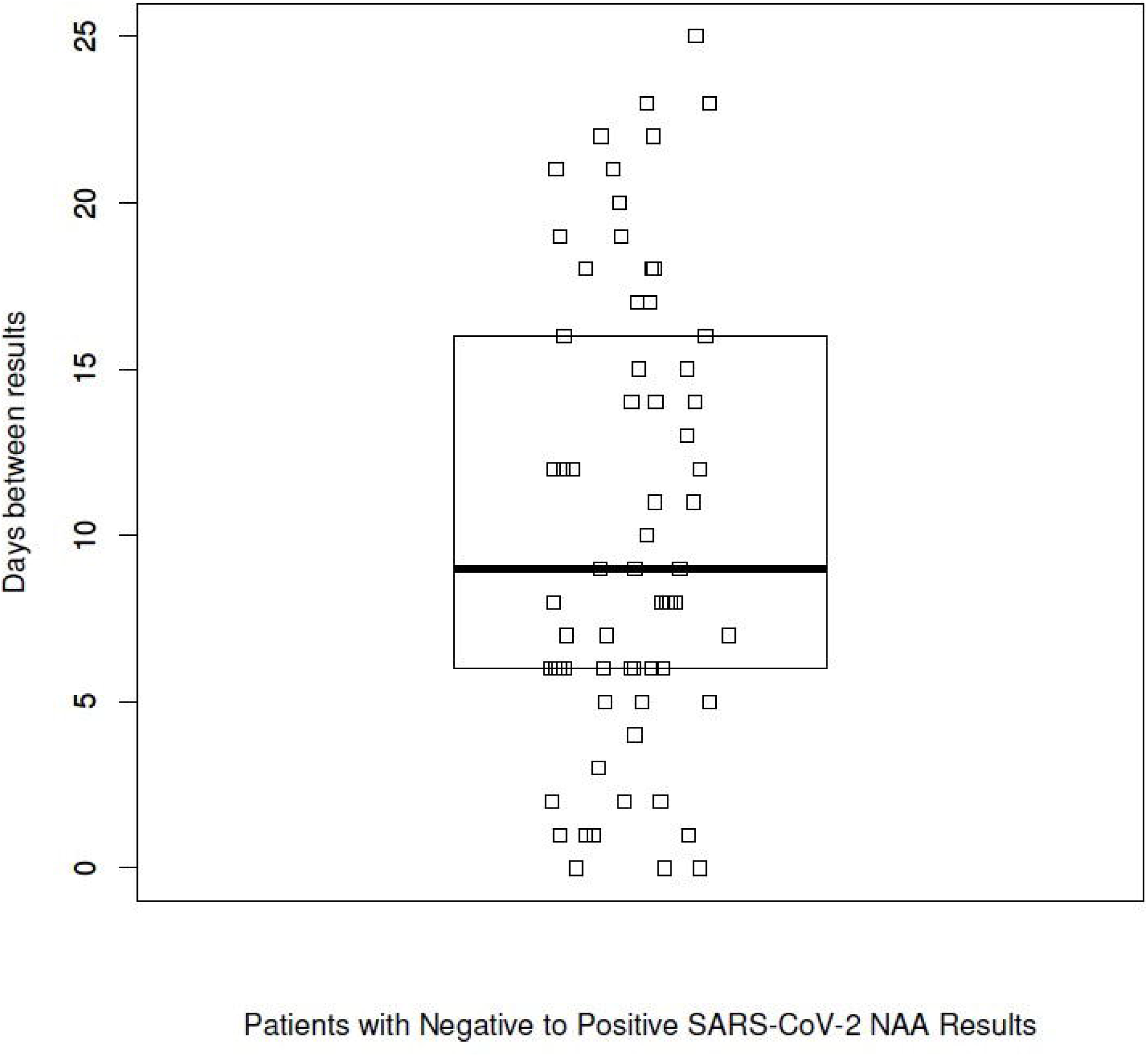
Days between progression (switch from negative to positive SARS-CoV-2 test results). Box shows interquartile range and median.

**Figure 3:**
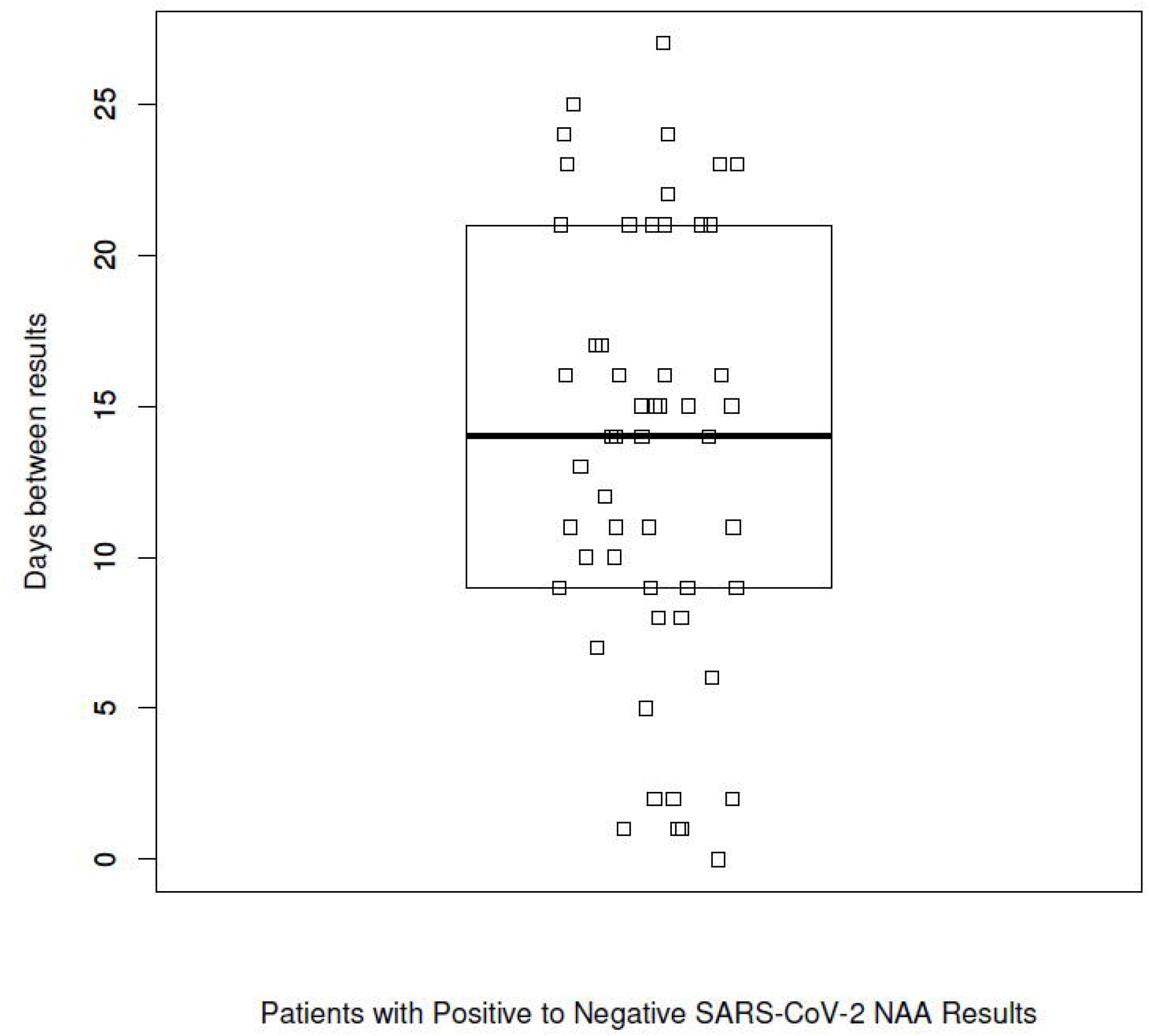
Days between remission (switch from positive to negative SARS-CoV-2results). Box shows interquartile range and median.

## CONCLUSION

Recent guidance from the Centers for Disease Control and Prevention (CDC) has been released regarding a Symptom-Based Strategy to Discontinue Isolation for Persons with COVID-19. ^11^ While the Symptom-Based strategy is appropriate for many patients, interim guidance from the CDC includes options for discontinuing isolation precautions for asymptomatic test-positive patients are divided into two different strategies, time-based and test-based. Isolation times for patients <u>who have never had any symptoms</u> using the time-based strategy require 10 days since the positive test. With the test-based strategy, asymptomatic carriers can be cleared from isolation with *two negative tests from different samples which are more than 24 hours apart* (see CDC guidance for details).^12^ These recommendations highlight both the variability in viral shedding and sampling errors possible in affecting testing results.

In our study, same day repeat testing was a small proportion of patients who had a change in test results (0 days to progression/remission). These few patients were likely due to differences in nasopharyngeal swabbing technique rather than progression from a carrier state or remission of viral production in recovering patients. It is likely that some of the patients with change in lab results in the 1-2 day time frame may also represent a change in test results due to nasopharyngeal swabbing technique. Repeat sample collection with nasal/nasopharyngeal swab in a brief window of time is recommended to confirm a negative test.^4,13^ Little information is available regarding false positive test results which could confound an early carrier/incubation/low symptom/convalescent carrier period in which patients lack symptoms and test positive. ^14^ However, divergent coronavirus strains or phylogenetically similar viral genomes could be involved in a repeated positive test in the absence of symptoms. ^6,13^ Presence of viral nucleic acid long after viral cultures can be recovered (i.e., culture serves as an in vitro indication of in vivo organism viability) has been demonstrated and could be contributing to difficulties clearing patients from isolation with a nucleic acid test-based strategy.^15,16^

Although knowledge about viral incubation times is rapidly evolving and many studies have highly varied results, the data collected here is concordant with what is currently known about viral incubation times and post-recovery carriage times. The “progression” data collected in this study overlaps with studies which show viral incubation time of between 2 and 14 days.^1,2,6,14,17^ The “remission” data collected in this study likewise suggests a carriage time following resolution of symptoms which extends for several weeks.^1,15,16^

The data presented here represents both clinical variability of sample collection as well as biological behavior of SARS-CoV-2 through the window of changes in laboratory testing. However, some recommendations on repeat testing can be made. For patients with high clinical suspicion and test results which are divergent from the clinical picture, swift repeat testing is recommended.^6,13^ If repeat testing is confirmatory, and an additional repeat test is required by the clinical circumstances, repeat testing after a period of time within the interquartile ranges of collected data, such as the median (Figures 2,3) should be considered. Therefore, testing for progression following a repeat negative is recommended at 9 days (or between 6 and 16 days) and testing for remission following a repeat positive is recommended at 14 days (or between 9 and 21 days).

In conclusion, although some guidance can be drawn from this data for repeat testing, this study does not control for sampling of the nares, testing methodology, patient disease state, or time of sampling. Long term follow-up on a broader window of patients is not available at the time of writing this and would give a more accurate time frame for repeat testing. Additionally, correlation with serologic testing could give this data both additional validity for recommending testing times and a deeper understanding of disease and immunologic dynamics. Finally, clinical details surrounding the repeat testing are unavailable to the authors and would give a more thorough understanding of effective utilization of testing for SARS-CoV-2.

## Data Availability

All data, blinded to study participant identifiers, is available to reviewers upon request.

